# Evaluating the Levels of Calprotectin in Patients with Chronic Non-specific Colitis: A Brief Report

**DOI:** 10.1101/2023.04.08.23288148

**Authors:** Rahmatollah Rafiei, Amin Najjar Khodabakhsh, Fereshteh Rafiei, Amirhossein Kamyab, Alireza Ebrahimi, Soheil Ashkani-Esfahani

## Abstract

**Background:** Patients with gastrointestinal symptoms of chronic diarrhea, chronic constipation, and abdominal pain might have unspecified signs and symptoms making them hardly characterized. These patients could be labeled as chronic nonspecific colitis patients. In this investigation, we aimed to compare the therapeutic effects of mesalamine in chronic nonspecific colitis patients with by measuring the levels of fecal calprotectin and assessing the reduction of their symptoms during the treatment.

**Methods:** Eighty-four outpatients (42 patients with normal, and 42 having high calprotectin levels) participated in this study. Participants were being treated by 2 grams of mesalamine for one month, and they were evaluated weekly. After one month, the participants’ signs and symptoms were reviewed, and the patients were labeled as treated or untreated.

**Results:** The result of this study demonstrated that there was no significant difference between the level of calprotectin among male or female participants, patients with different ages, or patients with different symptoms. Calprotectin levels were significantly different between patients who positively responded to mesalamine treatment compared to those who did not (p<0.001). On the other hand, patients with high calprotectin levels had a higher response rate to mesalamine (94.5%), compared to those with normal calprotectin levels (36.1%; p<0.001).

**Conclusion:** There was a significant correlation between the level of fecal calprotectin and the response rate of the patients to mesalamine; therefore, this parameter might be a good indicator to be used for treatment plans. Further studies are suggested to affirm this outcome.

## 1. Introduction

Non-specific colitis (NSC) refers to a condition in which clinical information is insufficient for a definitive diagnosis despite microscopic evidence of inflammation in the large intestine [1]. NSC patients sometimes present symptoms such as diarrhea, constipation, or abdominal pain [1]; however, they cannot be classified as inflammatory bowel disease (IBD) or irritable bowel syndrome (IBS) [2, 3]. The incidence of NSC is globally increasing; although its pathogenesis and treatment remain obscure, it is sometimes misdiagnosed and mistreated even after a thorough endoscopy and colonoscopy, leading to more severe conditions [2, 4, 5]. Therefore, finding a reliable biomarker in order to plan treatment for the disease remains a concern.

Beside radiological, macroscopic, and histological evaluations of the small bowel and colon, there are biochemical markers that can be used as indicators of gastrointestinal (GI) inflammation, for instance, erythrocyte sedimentation rate (ESR), C-reactive protein (CRP), and orosomucoid levels [6]. Fecal markers such as calprotectin and lactoferrin, have higher specificity for the diagnosis of GI diseases in comparison with serological markers [7-11]. Fecal calprotectin is used as a determinant for grading intestinal inflammation and predicting IBD flares [7-11]. It has been asserted that in contrast with ESR, CRP, or orosomucoid which cannot predict IBD flares, fecal calprotectin has about 70% sensitivity and specificity for prediction of IBD relapses [12].

In this investigation, we aimed to compare the response rate to treatment with mesalamine in NSC patients with high and normal levels of fecal calprotectin and assess the reduction of their symptoms during the treatment.

## 2. Methods

### 2.1. Study Design and Population

In this longitudinal study, 500 patients who were referred to the gastroenterology department of Shariati hospital, Isfahan, Iran, between October 2018 and October 2019, and were suspicious of inflammatory bowel disease according to colonoscopic findings were evaluated. Specimens obtained from IBD patients who had colonoscopy indication, and were examined by an expert pathologist. The diagnosis of NSC was based on microscopic evaluation of biopsies. The specimens showing the inflammation of the tissue but could not be classified as IBD were labeled as NSC [13, 14]. Fecal calprotectin levels were also measured in the participants. The cutoff point for the calprotectin level was specified ≥50 μg/g feces according to previously published studies [15]. In the end, 42 patients with normal calprotectin levels (calprotectin of <50 μg/g) and 42 individuals with high fecal calprotectin levels (calprotectin of ≥50 μg/g) were recruited and allocated into group 1 and group 2, respectively.

### 2.2. Inclusion and Exclusion Criteria

Inclusion criteria were clinical and pathological confirmation of NSC according to colonoscopic and pathologic findings, and not having used mesalamine in a month prior to the study. Exclusion criteria were the presence of rectorrhagia as it can disrupt the calprotectin test, a positive history of other confirmed GI diseases such as IBS, Celiac, GI food allergies, autoimmune GI disorders, chronic liver diseases, positive history of recent antibiotic consumption, alcohol abuse, smoking, occurrence of side effects during the treatment, and patients’ unwillingness to participate in the study.

### 2.3. Procedure

Patients with normal and high fecal calprotectin levels were being treated by 2 grams (1 gr, twice daily) of mesalamine (500 mg enteric-coated tablet, Arya Pharmaceutical Company, Tehran, Iran) for one month. Every two weeks (days 0, 15, and 30), physical examination of the participants was done by an expert gastroenterologist and a previously designed questionnaire evaluating their abdominal pain through a 10-point scale, frequency and intensity of symptoms, and their overall score for quality of life, was asked to fill. The patients were labeled as responsive or non-responsive to the treatment according to earlier published literature and based on Rome IV criteria [16].

### 2.4. Statistical Analysis

After data gathering, the results were analyzed using SPSS statistical software (ver. 18.0, IBM™, USA). The statistical analyses were carried out by employing independent t-test, and P < 0.05 was considered statistically significant.

### 2.5. Ethical Considerations

The study’s protocol was approved by the Medical Ethics Committee of Islamic Azad University of Najafabad, Isfahan, Iran (Reg. No: ir.iau.najafabad.rec.1396.79). Before the study, a written informed consent was obtained from all participants, and their anonymity was guaranteed.

## 3. Results

According to the results of this study, no significant difference was witnessed between the two groups of the study in terms of age and gender (Table-1).

As shown in Table 2, there were no significant difference between two groups regarding the frequency of gastrointestinal symptoms, including abdominal pain as the most predominant symptom, diarrhea, and constipation.

**Table 1.** Demographic data of patients diagnosed with non-specific colitis with normal (group 1, n=42) and high fecal calprotectin level (group 2, n=42). Data are expressed as mean (standard deviation).

| Variable | Group 1 | Group 2 | P-value |
| --- | --- | --- | --- |
| Age (years) | 45.88 ± 4.18 | 40.16 ± 14.4 | 0.07 |
| Gender (M/F) | 52.4%/47.6% | 40.5%/59.5% | 0.28 |

**Table 2.** The frequency of gastrointestinal symptoms in the two groups of the study.

| Symptoms | Group 1 | Group 2 | P-value |
| --- | --- | --- | --- |
| Abdominal pain | 85.7% | 88.1% | 0.9 |
| Diarrhea | 9.5% | 14.3% | 0.8 |
| Constipation | 2.4% | 0 | 0.9 |

According to the results, there was a significant difference between the two groups regarding their response to treatment with mesalamine. Patients having high fecal calprotectin levels (Group 2) showed better overall improvement compared to patients with normal calprotectin levels (Group 1) (p<0.001). However, regarding the symptoms, only abdominal pain showed significantly better relief in group 2 compared to group 1 (Table 3). No significant differences were seen regarding the response of diarrhea and constipation to treatment with mesalamine.

**Table 3.**
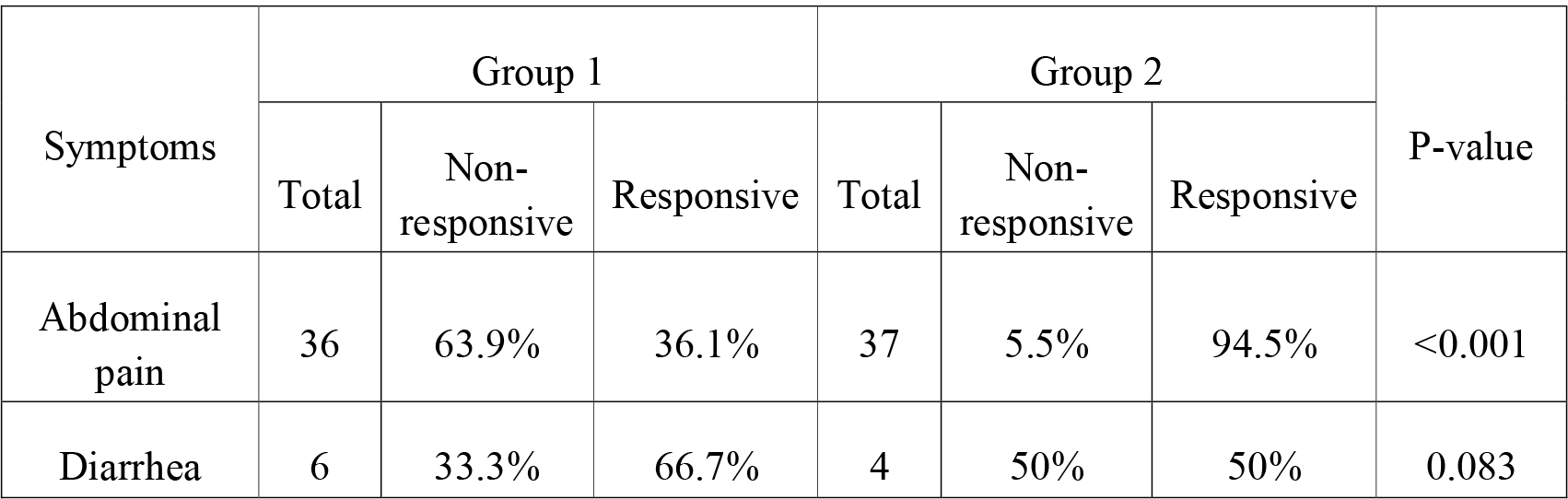
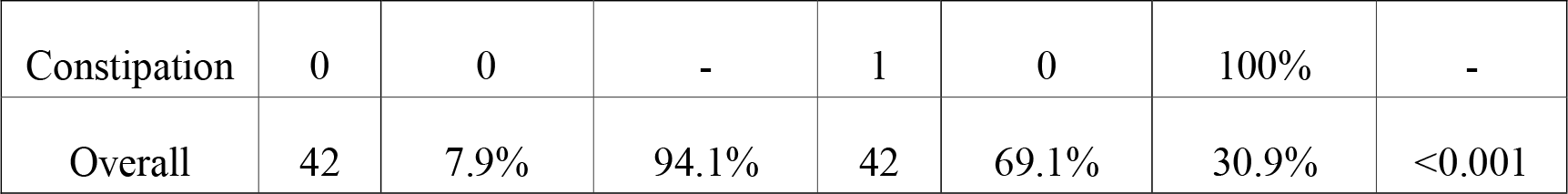
Response rate of patients diagnosed with non-specific colitis with normal (group 1, n=42) and high fecal calprotectin levels (group 2, n=42) treated with mesalamine.

| Symptoms | Group 1 |  |  | Group 2 |  |  | P-value |
| --- | --- | --- | --- | --- | --- | --- | --- |
|  | Total | Non-responsive | Responsive | Total | Non-responsive | Responsive |  |
| Abdominal pain | 36 | 63.9% | 36.1% | 37 | 5.5% | 94.5% | <0.001 |
| Diarrhea | 6 | 33.3% | 66.7% | 4 | 50% | 50% | 0.083 |
| Constipation | 0 | 0 | - | 1 | 0 | 100% | - |
| Overall | 42 | 7.9% | 94.1% | 42 | 69.1% | 30.9% | <0.001 |

## 4. Discussion

This study was designed to evaluate clinical symptoms of NSC patients having high and normal fecal calprotectin levels in response to treatment with mesalamine. To the best of our knowledge, no study have evaluated fecal calprotectin levels in patients who are being treated with the diagnosis of NSC; however, some investigations examined the biomarker in other diseases such as ulcerative colitis and colonic diverticular diseases [17, 18].

Our study showed NSC cases with higher fecal calprotectin levels who received mesalamine as the treatment has a better response rate compared to those with normal levels of calprotectin regarding their clinical symptoms. Previous studies demonstrated the same conclusion in patients with UC as fecal calprotectin found to be significantly lower in patients with clinical and endoscopic remission during mesalamine suppository treatment [17]. Researchers also suggested that the increment of mesalamine dose could decrease fecal calprotectin concentration in UC patients which could be because of a reduction in the tract inflammation [19]. Previous investigations asserted that fecal calprotectin is an indicator of treatment and a predictor of relapse in patients with bowel inflammation [20]. According to our results, this biomarker could also be used as an indicator to predict the response rate to the treatment in NSC patients. Husebye et al. reported that fecal calprotectin has negative predictive value in the case of colonic inflammation and neoplasm [21]. Moreover, fecal calprotectin was described as a viable tool to predict colonic inflammation in patients with chronic diarrhea [22].

The gold standard for diagnosis of NSC is an endoscopic or colonoscopic evaluation with histologic confirmation besides clinical examination. Additional markers such as ESR, CRP, and orsomucoid levels could also be beneficial in case of definite conclusions. Fecal calprotectin concentration assessment is perceived as a practical, non-invasive and cost-benefit tool for monitoring bowel inflammation [23, 24]. Considering the lack of literature with respect to using biomarkers in the treatment process of NSC patients our study is among the first papers discussing fecal calprotectin as a beneficial marker for monitoring the treatment process of these patients.

This study had several limitations to be considered. The concrete definition of NSC is yet to be cleared, as there is no specific GI tract structural deformity that describes the condition. Pathologists may label a specimen as NSC when they are not able to classify the pathological changes into a specific type of IBD. There are also probable methodological limitations for this study such as conducting biopsy on incorrect sites.

## 5. Conclusion

The present study has shown NSC patients who had high fecal calprotectin levels are more responsive to mesalamine treatment. Abdominal pain was the most abundant symptom among NSC patients and patients with high fecal calprotectin levels showed higher relief rate compared to those with normal calprotectin levels. Further studies are required to establish the cost-benefit and precision of using fecal calprotectin in as a predictor in the treatment of patients suffering from IBD.

## Data Availability

The datasets used and/or analyzed during the current study are publicly available from the corresponding author on reasonable request.

